# Bridging AI and Healthcare: A Scoping Review of Retrieval-Augmented Generation—Ethics, Bias, Transparency, Improvements, and Applications

**DOI:** 10.1101/2025.04.01.25325033

**Authors:** David J. Bunnell, Mary J. Bondy, Lucy M. Fromtling, Emilie Ludeman, Krishnaj Gourab

**Author notes:** Corresponding author – David J. Bunnell.

## Abstract

**Background:** Retrieval-augmented generation (RAG) is an emerging artificial intelligence (AI) strategy that integrates encoded model knowledge with external data sources to enhance accuracy, transparency, and reliability. Unlike traditional large language models (LLMs), which are limited by static training data and potential misinformation, RAG dynamically retrieves and integrates relevant medical literature, clinical guidelines, and real-time data. Given the rapid adoption of AI in healthcare, this scoping review aims to systematically map the current applications, implementation challenges, and research gaps related to RAG in health professions.

**Methods:** A scoping review was conducted following the Joanna Briggs Institute (JBI) framework and reported using the Preferred Reporting Items for Systematic Reviews and Meta-Analyses for Scoping Reviews (PRISMA-ScR) guidelines. A systematic search strategy was designed in collaboration with faculty and a research and education librarian to include PubMed, Scopus, Embase, Google Scholar, and Trip, covering studies published between January 2020 and August 2024. Eligible studies examined the use of RAG in healthcare. Studies were screened in two stages: title/abstract review followed by full-text assessment. Data extraction focused on study characteristics, applications of RAG, ethical and technical challenges, and proposed improvements.

**Results:** A total of 31 studies met inclusion criteria, with 90.32% published in 2024. Authors came from 17 countries with the most frequent publications coming from the USA (*n* = 15), China (*n* = 3), and the Republic of Korea (*n* = 3). Key applications included clinical decision support, healthcare education, and pharmacovigilance. Ethical concerns centered on data privacy, algorithmic bias, explainability, and potential overreliance on AI-generated recommendations. Bias mitigation strategies included dataset diversification, fine-tuning techniques, and expert oversight. Transparency measures such as structured citations, traceable information retrieval, and explainable diagnostic pathways were explored to enhance clinician trust in AI-generated outputs. Identified challenges included optimizing retrieval mechanisms, improving real-time integration, and standardizing validation frameworks.

**Conclusion:** RAG AI has the potential to improve clinical decision-making and healthcare education by addressing key limitations of traditional LLMs. However, significant challenges remain regarding ethical implementation, model reliability, and regulatory oversight. Future research should prioritize refining retrieval accuracy, strengthening bias mitigation strategies, and establishing standardized evaluation metrics. Responsible deployment of RAG-based systems requires interdisciplinary collaboration between AI researchers, clinicians, and policymakers to ensure ethical, transparent, and effective integration into healthcare workflows.

## Introduction

Artificial intelligence (AI) is being explored in healthcare for clinical decision support, diagnosis, and patient engagement. However, traditional large language models (LLMs) face limitations, including reliance on static model training data, susceptibility to outdated information, and generating unverifiable or misleading content (Amugongo et al., 2024; Ng et al., 2025). Despite performing well on medical licensing exams, large language models perform worse than clinicians in real-world clinical decision-making tasks (Hager et al., 2024). Retrieval-augmented generation (RAG) addresses these challenges by combining a model’s neural network-based knowledge with external retrieval from structured sources, including medical literature, clinical guidelines, and databases (Lewis et al., 2020). This approach enhances accuracy, transparency, and relevance, mitigating AI-generated misinformation and improving explainability (Amugongo et al., 2024).

Despite its promise, RAG’s applications in healthcare remain underexplored (Ng et al., 2025). While initial research suggests its potential to improve real-time clinical decision-making and evidence-based recommendations, challenges persist. These include a lack of standardized evaluation frameworks, ethical concerns related to privacy and bias, and the complexity of optimizing retrieval mechanisms to minimize irrelevant or misleading information (Amugongo et al., 2024). A recent systematic review emphasizes the need to refine retrieval strategies, integrate multimodal data, and develop robust validation methodologies to ensure RAG enhances clinical workflows without introducing additional risks (Amugongo et al., 2024).

Given the rapid evolution of RAG and its potential to transform healthcare, a scoping review is necessary to systematically map its current applications, assess implementation challenges, and identify gaps in the literature. This review specifically provides insights into RAG strategies to augment LLMs from the clinician’s point of view. This will inform future research and guide the responsible integration of RAG-based systems in healthcare settings.

## Methods

This scoping review was conducted following the methodological framework outlined by the Joanna Briggs Institute (JBI) and reported in accordance with the Preferred Reporting Items for Systematic Reviews and Meta-Analyses for Scoping Reviews (PRISMA-ScR) (see Appendix A) (Peters et al., 2020). Studies were included if they focused on health professions and explored the use of RAG within healthcare contexts (see Table 1). Study designs that were eligible included randomized controlled trials, quasi-experimental studies, observational studies, qualitative studies, comparative effectiveness, and mixed-methods research. Eligible publication types included peer-reviewed journal articles, preprints, dissertations, and theses. Only studies published in English were considered.

**Table 1.** Inclusion and Exclusion Criteria.

| Inclusion | Exclusion |
| --- | --- |
| **Population:** Studies involving health professions. | **Population:** Studies not related to health professions |
| **Intervention:** Studies focusing on the use of Retrieval Augmented Generation (RAG) in all contexts. | **Intervention:** Studies not involving Retrieval Augmented Generation (RAG) as a primary focus. |
| **Study Design:** All study designs, including randomized controlled trials, quasi-experimental studies, observational studies, qualitative studies, and mixed-methods studies. |  |
| **Outcomes:** Studies reporting on applications, implementation strategies, effectiveness, benefits, challenges, gaps, or future research needs related to RAG in health professions. | **Outcomes:** Studies that do not report on applications, implementation strategies, effectiveness, benefits, challenges, gaps, or future research related to RAG in health professions. |
| **Publication Type:** Peer-reviewed journal articles, preprint articles, dissertations, and theses. | **Publication Type:** Opinion pieces, editorials, conference papers, and news articles. |
| **Language:** Publications in English. | **Language:** Publications not available in English. |
| **Publication Date:** 2020 to August 19, 2024 | **Publication Date:** Studies published after the search cut-off date. |

Exclusion criteria were applied to filter out studies that did not pertain to health professions, or did not focus primarily on RAG (see Table 1). Opinion pieces, editorials, conference papers, and news articles were also excluded. The search was conducted on August 19, 2024 and limited to articles published since 2020, aligning with the introduction of RAG by Lewis et al (2020).

A systematic search of the literature was performed by a research and education librarian (EL) across multiple databases, including PubMed (*n* = 84), Scopus (*n* = 37), Embase (*n* = 35), Google Scholar (*n* = 14), and Trip (*n* = 6). The search strategy incorporated a combination of RAG-related terms and health professions-related terms (see Table 2). Key terms included variations of retrieval-augmented generation, retrieval-augmented text generation, and retrieval-augmented language models, as well as health professions-related terms such as health professions education, medicine, nursing, pharmacy, and dentistry. Boolean operators and controlled vocabulary (e.g., MeSH terms) were applied where appropriate.

**Table 2.** Search Strategy for Identifying Articles on Retrieval-Augmented Generation.

| Concept | Search Terms (Field Tags) |
| --- | --- |
| Retrieval-Augmented Generation (RAG) | "retrieval augmented generation"[tiab]<br>"retrieval augmented text generation"[tiab]<br>"generation augmented retrieval"[tiab]<br>"retrieval augmented controllable review generation"[tiab]<br>"retrieval augmented code generation"[tiab]<br>"retrieval augmented model"[tiab]<br>"retrieval augmented language models"[tiab]<br>"augment* generative models"[tiab] |
| Health Professions and Medical Education Contexts | HPE[tw]<br>health professions education[tw]<br>medical[tw]<br>medicine[tw]<br>nursing[tw]<br>pharmacy[tw]<br>dental[tw]<br>dentistry[tw] |
| Boolean Logic Applied | (RAG terms) AND (HPE terms) |

Field Tags: [tiab] = Title/Abstract, [tw] = Text Words (includes title, abstract, MeSH terms, and other fields)

Study selection followed a two-step process. First, titles and abstracts were screened independently by two reviewers (DJB and MJB) to identify eligible studies. Those meeting the inclusion criteria proceeded to full-text review, conducted by three reviewers (DJB, LMF, and MJB). Any discrepancies were resolved through discussion when necessary.

To ensure data extraction consistency, a standardized data charting form was developed and piloted in the Covidence systematic review software (see Appendix B). Reviewers conducted calibration exercises prior to data extraction to enhance reliability (DJB and LMF). Data extracted included study characteristics, RAG applications, and ethical and technical challenges associated with RAG in healthcare. No formal critical appraisal of the included studies was conducted. Instead, the synthesis focused on providing a comprehensive overview of the available literature. Data were synthesized descriptively and thematically using OpenAI’s ChatGPT-4o with review and confirmation by the researchers. This qualitative data was reviewed for the following categories: ethical issues, bias mitigation strategies, transparency and explainability, proposed improvements, and potential model applications.

## Results

A total of 128 records were screened for inclusion in this scoping review. Of these, 97 records were excluded, including 71 based on abstract review and 26 following full-text assessment (see Figure 1). The reasons for exclusion during the full-text review were non-research articles such as conference proceedings (*n* = 9), opinion pieces, editorials, or essays (*n* = 7), and studies for which the full text was unavailable (*n* = 7). Additional exclusions included articles with an inappropriate study design (*n* = 2) and one duplicate record. Study designs identified as inappropriate for this study included one utilizing non-RAG strategies and another reporting results from a workshop. Ultimately, 31 studies met the inclusion criteria and were incorporated into the final analysis.

**Figure 1.**
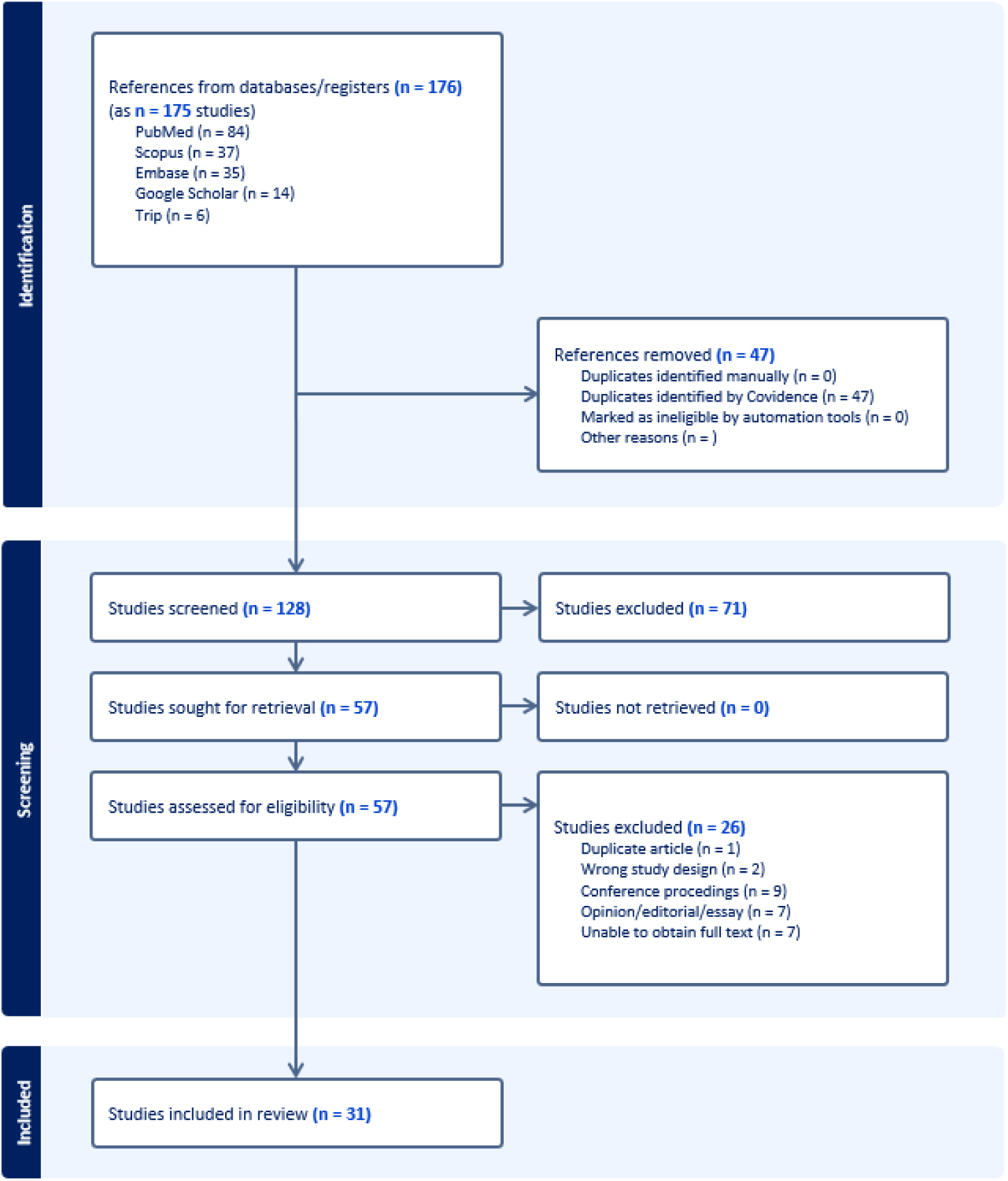
PRISMA Diagram.

Data was extracted from 31 publications, with 28 (90.32%) published in 2024 and only 3 (9.68%) in 2023 (see Figure 2). Regarding peer review status, 12 (38.71%) of the publications were peer-reviewed, while 19 (61.29%) were not peer-reviewed. Funding distribution was nearly balanced, with 16 (51.61%) studies reporting funding and 15 (48.39%) not reporting any funding.

**Figure 2.**
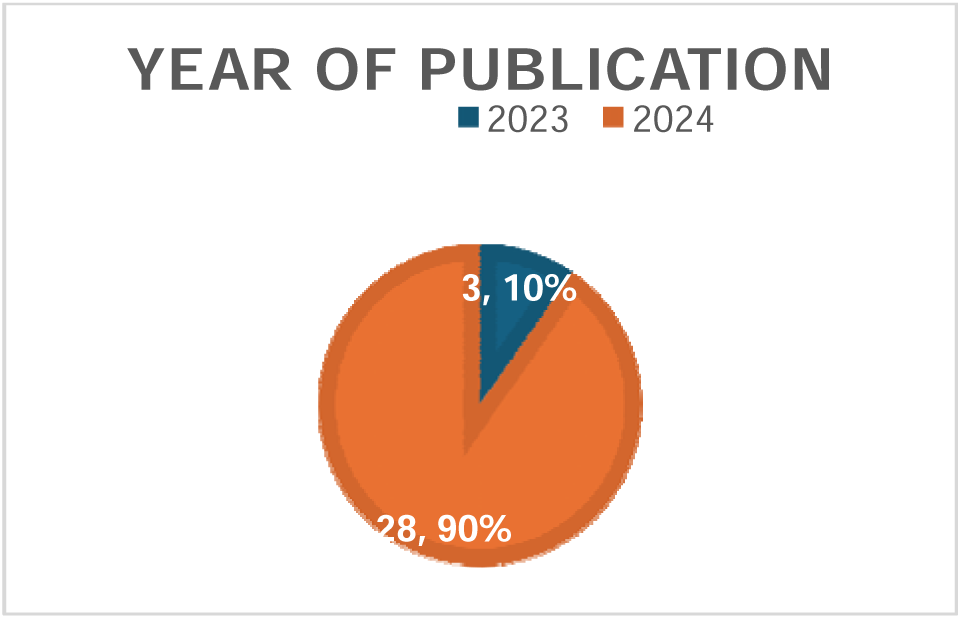
Year of Publication.

The included studies represent research from 17 different countries, with the United States (*n* = 15) contributing the highest number of publications (see Table 1 and Figure 3). Other countries with multiple publications include China (*n* = 3), the Republic of Korea (*n* = 3), Australia (*n* = 2), and Italy (*n* = 2). The remaining countries—Denmark, Ghana, India, Iran, Saudi Arabia, Singapore, Spain, Sweden, Taiwan, Turkey, and the United Kingdom—each contributed one publication.

**Table 3.** Country Identified by Authors.

| Country | Number of Publications |
| --- | --- |
| Australia | 2 |
| China | 3 |
| Denmark | 1 |
| Ghana | 1 |
| India | 1 |
| Iran | 1 |
| Italy | 2 |
| Republic of Korea | 3 |
| Saudi Arabia | 1 |
| Singapore | 1 |
| Spain | 1 |
| Sweden | 1 |
| Taiwan | 1 |
| Turkey | 1 |
| UK | 1 |
| USA | 15 |

**Figure 3.**
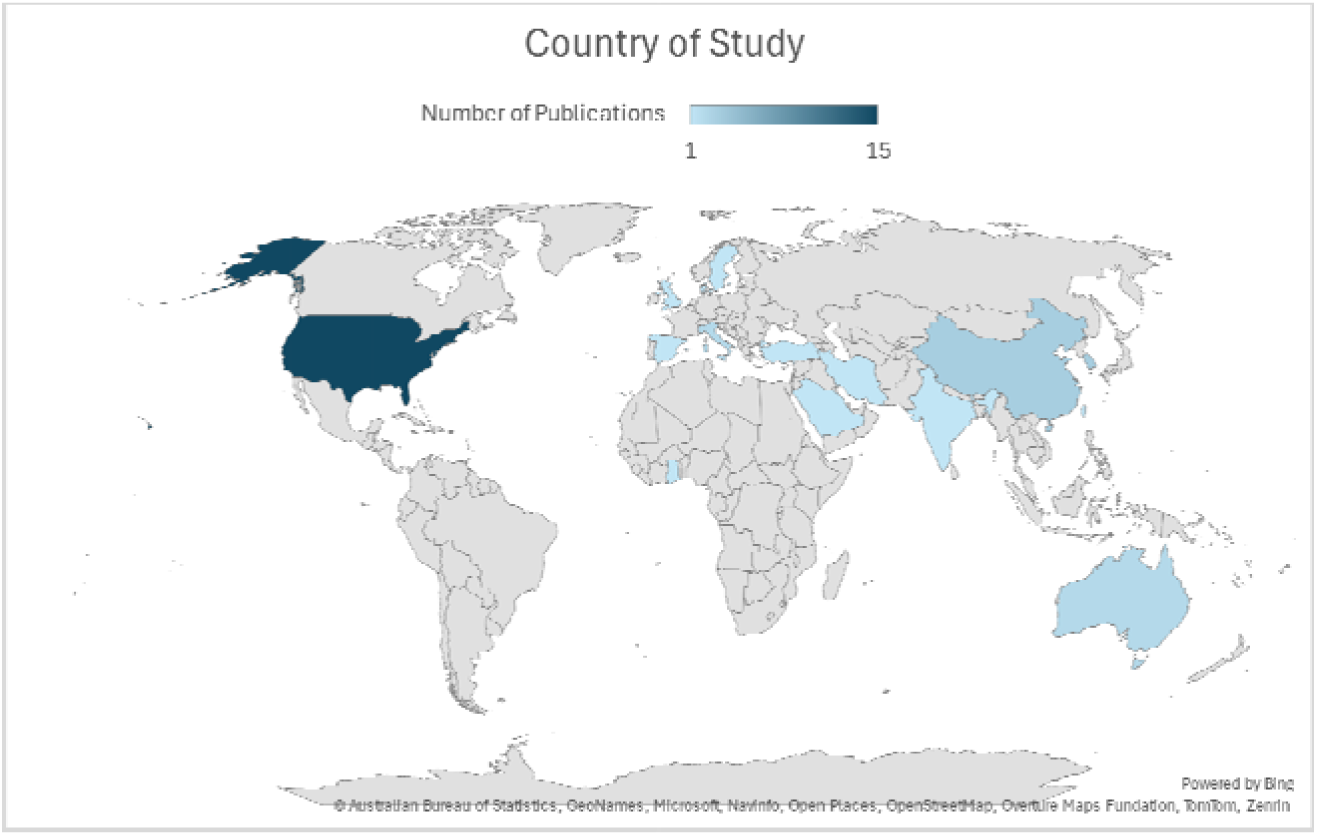
Country of Study.

### Ethical Issues Raised

The scoping review identified multiple ethical considerations for using RAG strategies in healthcare applications. These considerations were grouped into six themes: privacy and data security, bias and cultural sensitivity, transparency and explainability, overreliance on AI in decision-making, ethical accountability and safeguards, and contextual appropriateness in resource-constrained settings.

Privacy and Data Security were identified as primary concerns. RAG enhanced LLM conversations are often stored, raising issues regarding data usage, security, and potential privacy breaches (Tegsten, 2024). The risk of data exposure was highlighted, with concerns that service providers could share or utilize sensitive medical information (AlGhadban et al., 2023; Tegsten, 2024). AI models’ reliance on patient data was noted as a contributing factor to increased cybersecurity risks (Quidwai & Lagana, 2024; Tegsten, 2024). Ensuring patient data anonymization and compliance with privacy regulations was emphasized as a necessary safeguard (Quidwai & Lagana, 2024).

The potential for bias and the need for cultural sensitivity were discussed. Studies highlighted the risk of algorithmic bias influencing clinical recommendations (Ge et al., 2024). Concerns were raised regarding ensuring that RAG enhanced LLM strategies fairly represent diverse cultural and medical practices, including traditional medicine approaches such as Traditional Chinese Medicine (Su & Gu, 2024). The risk of bias in responses was further compounded by limitations in training data and the potential for AI models to reinforce existing disparities (AlGhadban et al., 2023; Long et al., 2024). Additionally, the importance of avoiding controversial or ethically sensitive topics, such as predicting fetal gender, was noted as a necessary precaution in AI-driven healthcare applications (AlGhadban et al., 2023).

Transparency and explainability were identified as key ethical concerns. Studies highlighted the opaque nature of general LLMs, noting that a lack of transparency poses risks in medical applications. The issue of explainability was discussed to ensure clinicians could understand and trust AI-generated responses (Chen et al., 2024). Algorithmic transparency and accountability were emphasized as essential for the responsible implementation of AI in healthcare (Gao et al., 2023; Tegsten, 2024; Wang et al., 2024).

The risk of overreliance on AI for medical decision-making was another recurring concern (Gao et al., 2023; Ke et al., 2024). LLMs were identified as being susceptible to hallucinations and positional bias in responses, which could lead to inaccurate or misleading medical guidance (Liévin et al., 2024). The potential for AI-generated misinformation was noted, mainly when responses were based on non-peer-reviewed sources (Wang et al., 2024). Studies also highlighted the need for strong human oversight when using LLMs for high-risk clinical decision-making to mitigate potential harms (Ke et al., 2024).

Ethical accountability and safeguards were emphasized in discussions about AI deployment in healthcare. The need for robust safeguards to prevent harmful content creation was identified as a critical issue (Liévin et al., 2024). Several studies highlighted concerns about deploying RAG enhanced LLMs without adequate safety mechanisms, mainly due to their susceptibility to generating misleading information (Wang et al., 2024). Strategies such as local data storage, transparent AI development practices, and dynamic data integration without compromising privacy were cited for addressing ethical accountability (Tegsten, 2024).

Finally, contextual appropriateness in resource-constrained settings was identified as a consideration in implementing RAG enhanced LLMs in healthcare. Ensuring that AI-generated outputs align with local healthcare practices was emphasized (Mensah et al., 2024). Mensah et al. (2024) noted that LLM-generated recommendations could be unhelpful or potentially harmful if they failed to consider the constraints of specific healthcare environments.

### Bias Mitigation Strategies

The studies identified multiple approaches to addressing bias, categorized into six key areas: data diversity and representation, fine-tuning and embedding methods, evaluation and expert oversight, contextual and ethical safeguards, prevention of hallucinations and misinformation, and ethical and social considerations.

RAG models leverage known and accurate data which aims to improve LLM responses. Studies incorporated diverse datasets to enhance fairness and reduce bias (Alghamdi & Mostafa, 2024; Ge et al., 2024; Liévin et al., 2024; Long et al., 2024; Su & Gu, 2024; Tegsten, 2024; Wang et al., 2024). Other studies emphasized the integration of reputable academic sources and included diverse and international guidelines to ensure broad representation (Ke et al., 2024; Wang et al., 2024).

Fine-tuning strategies were applied to optimize AI models for healthcare-specific applications (Gao et al., 2023; Liévin et al., 2024; Su & Gu, 2024). Studies reported using external retrievals, domain-specific prompts, and embeddings to improve biomedical context alignment (Kang et al., 2024; Su & Gu, 2024; Tegsten, 2024). Iterative evaluations were also conducted to refine model accuracy and response quality (Su & Gu, 2024).

Expert oversight and structured evaluation further supported bias mitigation. Cross-validation through clinician review verified the accuracy and consistency of AI-generated outputs (Wang et al., 2024). One strategy incorporated expert feedback to improve data labeling, while another applied aggregated scoring methods to reduce evaluator bias by averaging scores across multiple graders (Li et al., 2024; Upadhyaya et al., 2024). Structured formatting and oversight mechanisms were also employed to ensure systematic bias assessment (Kresevic et al., 2024).

The following strategies focused on ensuring cultural and contextual relevance in AI-generated content were discussed. Context-specific prompt engineering and domain-relevant resources, such as the Basic First Aid for Africa manual, aligned responses with regional and population-specific needs (Mensah et al., 2024). A model was described that included bias evaluation checks and underwent testing across multiple benchmarks to identify and address reasoning and response generation biases (Liévin et al., 2024).

Efforts to prevent hallucinations and misinformation were also documented. Implementing threshold-based retrieval mechanisms, where responses were generated only when sufficiently relevant information was available was demonstrated (Quidwai & Lagana, 2024). Quidwai and Lagana (2024) described that when data was insufficient, their model returned a response stating, “Sorry, I could not find relevant information to complete your request.” High-quality, domain-specific biomedical retrievers were also used to reduce generalization errors and enhance reliability (Jeong et al., 2024).

Ethical and social considerations were addressed through various design features. A study incorporated cultural sensitivity and impartiality measures while avoiding stereotypes in AI-generated responses (Alghamdi & Mostafa, 2024). Another model supported multi-language capabilities, enabling responses in Spanish and other languages to improve accessibility (Murugan et al., 2024). Additionally, safeguards were implemented to prevent AI-generated medical advice for patients, encouraging professional consultation for clinical decision-making (Murugan et al., 2024).

### Transparency and Explainability

Across the included studies, traceability of information retrieval was a focus, with systems implementing structured approaches to ensure that responses were linked to identifiable sources. Methods such as Triple Graph Construction and Graph-RAG were used to enhance reasoning and ensure that responses could be traced back to their sources (DiMaria et al., 2024; Wu et al., 2024). Systems also integrated hierarchical graphs to improve transparency in task decomposition, and guideline reformatting was noted as a mechanism to enhance traceability in model outputs (Kresevic et al., 2024; Long et al., 2024).

Citations and source documentation were key features of transparency-focused models (DiMaria et al., 2024; Li et al., 2024; Murugan et al., 2024; Wang et al., 2024). Additionally, user-accessible logs were provided, allowing users to review which external sources and LLM modules contributed to generated responses (Ke et al., 2024; Wang et al., 2024). Two models retrieved specific articles and passages to support responses, while citation metrics, impact factors, and metadata were integrated to improve transparency (Li et al., 2024; Su & Gu, 2024).

Structured and annotated approaches were used to improve model interpretability. This strategy for prompt engineering was incorporated to enhance explainability, while reflective tokens and evidence prioritization mechanisms were introduced to refine model outputs (Jeong et al., 2024; Upadhyaya et al., 2024). One model also employed explainable diagnostic pathways, ensuring that reasoning steps were accessible and structured (Gao et al., 2023).

Studies documented evaluation and accountability measures. Models included documentation of data preprocessing, integration processes, and evaluation metrics, such as confidence scores and sentiment analysis (Liu et al., 2024; Su & Gu, 2024). Additionally, disclaimers about AI limitations were provided. Some approaches included rationale explanations alongside responses to enhance interpretability (Murugan et al., 2024).

### Proposed Improvements

The scoping review identified a broad range of proposed improvements for RAG enhanced LLM strategies in healthcare, with key areas for enhancement including retrieval optimization, real-time integration, model fine-tuning, user interaction, benchmarking, and addressing AI challenges. Studies proposed improving retrieval techniques through neural retrieval models, semantic embeddings, and cross-encoder re-rankers (Su & Gu, 2024; Ye, 2024). Expanding data sources was suggested, including PubMed Central, FAQs from trusted sources, and biomedical QA corpora (Xiong et al., 2024). Additionally, multilingual retrieval mechanisms were identified as an area for development (Alonso et al., 2024).

Enhancements in real-time clinical data integration and AI optimization were proposed to improve clinical decision-making. Studies emphasized integrating real-time clinical data, continuously updating knowledge bases, and reducing response latency and computational costs (Long et al., 2024). Alternative RAG system architectures and enhanced embedding techniques were suggested to refine retrieval performance and efficiency (Tegsten, 2024; Ye, 2024).

Model fine-tuning and multimodal integration were key areas for improving LLM performance in healthcare applications. Fine-tuning LLMs using domain-specific datasets was a common recommendation (Alkhalaf et al., 2024; Alonso et al., 2024; Liévin et al., 2024; Long et al., 2024; Wang et al., 2024). Incorporating multimodal capabilities, such as computer vision for medical imaging and automatic speech recognition for clinical conversations, was suggested to expand AI applications in healthcare (Long et al., 2024). Bias mitigation strategies, domain-specific instruction sets, and de-identified patient data integration from electronic health record (EHR) systems were also identified as potential areas for improvement (Ge et al., 2024; Jeong et al., 2024; Quidwai & Lagana, 2024).

Improving user interaction and safety features was a recurring theme in the reviewed studies. Proposed improvements included structured prompt engineering techniques like Chain-of-Thought reasoning and real-time user feedback integration for iterative model refinement (Alghamdi & Mostafa, 2024; Long et al., 2024; Upadhyaya et al., 2024). Expanding chatbot functionalities to provide personalized recommendations and context-aware conversations was suggested (Alghamdi & Mostafa, 2024). Enhancing patient-friendly language and enriching knowledge bases to improve communication was also recommended (Murugan et al., 2024).

Benchmarking and performance evaluation were emphasized as essential components of AI system improvement. Future efforts were identified in developing systematic evaluation benchmarks, refining retrieval input constraints, and addressing token limitations (Jeong et al., 2024; Li et al., 2024). Suggested performance metrics included F1 scoring (a measure of a model’s accuracy), recall, and factual correctness, emphasizing improving model reliability through structured evaluation frameworks (Gao et al., 2023; Kresevic et al., 2024; Mensah et al., 2024; Ye, 2024).

Addressing AI challenges, including hallucinations, unpredictable variations in AI responses, and bias, was a focus across multiple sources. They proposed minimizing uncertainty in diagnostic processes, evaluating the correlation between automated metrics and human scoring, and refining fact-checking mechanisms to assess the neutrality of retrieved sources (Gao et al., 2023; Kresevic et al., 2024). Expanding label granularity and transitioning to deep learning-based classification were also suggested (Ye, 2024).

Advancements in open-source model development and collaboration were discussed as strategies for improving AI performance. Proposed advancements in open-source AI models included optimizing fine-tuning processes, expanding model parameter sizes, and integrating specialized medical databases (Alkhalaf et al., 2024; Liévin et al., 2024). Collaboration with healthcare professionals was suggested to enhance AI outputs, and structured stepwise feedback mechanisms were proposed for refining reasoning processes (Alghamdi & Mostafa, 2024; Upadhyaya et al., 2024).

### Potential Model Applications

The scoping review identified multiple potential applications for RAG AI in healthcare, clinical decision-making, and education. Studies described its possible use in patient education and public health monitoring, where it could track emerging drug side effects, assess the co-use of multiple substances, and analyze public perceptions of specific drugs (Das et al., 2024; Tegsten, 2024). The RAG AI strategy was also noted for its ability to conduct date-specific queries, enabling time-sensitive insights within selected timeframes (Das et al., 2024).

Applications in surgical and preoperative settings were identified, referencing using RAG AI for preoperative optimization and broader assessments of surgical workflows (Ke et al., 2024). The reviewed literature also highlighted its potential role in pharmacovigilance and drug safety, particularly in synthesizing information based on provided text to detect misinformation about specific substances (Das et al., 2024).

The potential for RAG AI to support clinical decision-making and precision medicine was noted, with studies indicating its possible integration into clinical decision-support systems and health informatics to provide real-time assistance (Alonso et al., 2024; Jeong et al., 2024; Long et al., 2024; Mensah et al., 2024; Wang et al., 2024). Additional applications were reported in developing decision support tools tailored for resource-constrained healthcare environments, with further consideration of its use in various domains of precision medicine (AlGhadban et al., 2023; Mensah et al., 2024; Quidwai & Lagana, 2024).

Several studies referenced the role of RAG AI in health professions education, including its potential to scale educational initiatives and support non-traditional learning environments (AlGhadban et al., 2023; Alonso et al., 2024; Choi et al., 2024; Wang et al., 2024). Specific applications were identified in ophthalmology diagnostics and the analysis of multimodal patient data, suggesting broader opportunities for integrating RAG AI into medical training and diagnostic processes (Upadhyaya et al., 2024).

## Discussion

This scoping review provides an overview of the current landscape of RAG AI models in healthcare, emphasizing ethical considerations, bias mitigation strategies, model transparency, proposed improvements, and potential applications. The findings highlight the rapidly evolving nature of RAG AI research, with most publications emerging in 2024. Despite the increasing interest in this domain, several challenges must be addressed to ensure the responsible and effective integration of RAG AI into clinical practice and health professions education.

### Ethical Considerations in RAG AI Implementation

A key finding from this review is the prominence of ethical concerns related to deploying RAG AI in healthcare settings. Privacy and data security remain paramount, with studies emphasizing the risks associated with LLM-generated conversations being stored, shared, or misused. Compliance with existing privacy regulations and the implementation of robust anonymization techniques are necessary to mitigate these risks. Additionally, bias and cultural sensitivity pose significant challenges, as AI models trained on skewed datasets may inadvertently reinforce healthcare disparities. The importance of ensuring fair representation of diverse medical practices and research was emphasized, particularly in studies addressing culturally specific treatments such as Traditional Chinese Medicine.

Due to the opaque nature of many AI models, transparency and explainability also emerged as ethical concerns. Clinicians must be able to trust and interpret AI-generated outputs, underscoring the need for improved algorithmic transparency and enhanced explainability in AI-driven healthcare applications. Furthermore, overreliance on AI for clinical decision-making raises concerns about misinformation, hallucinations, and the risk of clinicians placing undue trust in AI-generated responses. This necessitates the development of AI-assisted decision support systems with built-in validation mechanisms to ensure human oversight and minimize potential harm.

### Bias Mitigation Strategies and Transparency

Addressing bias in RAG AI remains crucial to ensuring equity in healthcare applications. This review identified multiple approaches to bias mitigation, including dataset diversification, fine-tuning methods, and expert oversight. While efforts to improve fairness through data diversity and domain-specific prompt engineering were reported, challenges persist in ensuring that AI-generated responses do not inadvertently reinforce existing biases. Additionally, structured evaluation methods and human oversight were cited as necessary safeguards to enhance the reliability of AI-generated medical recommendations.

The issue of transparency was explored in several studies, with various methodologies proposed to enhance model interpretability. Traceability mechanisms, such as Graph-RAG and structured citations, were employed to improve response verifiability. The integration of user-accessible logs and external source documentation also contributed to enhancing model transparency. However, ongoing research is needed to refine these approaches and standardize methods for ensuring accountability in AI-generated outputs.

### Proposed Improvements

The studies reviewed suggested numerous improvements to optimize the performance and reliability of RAG AI models in healthcare. Enhancing retrieval optimization, real-time data integration, and model fine-tuning were among the most frequently cited recommendations. The potential for integrating multimodal capabilities, such as combining AI-generated text with medical imaging and speech recognition, presents promising avenues for expanding the scope of AI applications in clinical practice.

User interaction improvements, including structured prompt engineering and real-time feedback mechanisms, were also proposed to refine AI-generated responses. Expanding patient-friendly language models and incorporating multi-language support could further enhance the accessibility of AI-driven healthcare tools. Additionally, benchmarking efforts, including developing systematic evaluation metrics such as F1 scores and recall rates, were emphasized as essential for ensuring model reliability.

### Potential Applications in Healthcare and Education

The findings highlight various potential applications for RAG AI in healthcare, spanning clinical decision support, patient education, and public health monitoring. The ability of AI models to track emerging drug side effects and analyze public perceptions of medical interventions underscores their potential in pharmacovigilance. Similarly, AI-driven preoperative optimization and surgical workflow assessments were identified as promising areas for RAG AI integration.

In clinical decision-making, RAG AI models have the potential to enhance precision medicine by providing real-time, evidence-based recommendations tailored to individual patient cases. However, careful validation and regulatory oversight remain critical considerations. The role of RAG AI in health professions education was also emphasized, with studies highlighting its ability to support medical training through personalized learning pathways and multimodal diagnostic tools.

### Limitations and Challenges

While this review comprehensively assesses the current literature on RAG AI in healthcare, several limitations must be acknowledged. The predominance of non-peer-reviewed sources highlights the need for further validation of study findings. Additionally, the rapid evolution of AI technologies means that ongoing research is necessary to stay abreast of new developments and emerging best practices. While bias mitigation strategies were widely discussed, their effectiveness in real-world clinical applications remains uncertain. Further empirical studies are required to assess the impact of these strategies on patient outcomes and healthcare equity. Ethical and regulatory challenges persist, particularly regarding data security, accountability, and the potential for AI-generated misinformation.

## Conclusion

This scoping review underscores the potential of RAG AI in healthcare, identifying critical ethical, technical, and regulatory challenges that must be addressed to ensure its responsible deployment. Bias mitigation, transparency, and enhanced user interaction remain key areas for future research. As AI models continue to evolve, interdisciplinary collaboration between healthcare professionals, AI researchers, and policymakers will be essential to maximize the benefits of RAG AI while minimizing risks. Future research should focus on refining model transparency, improving real-world validation methodologies, and developing standardized evaluation metrics to ensure AI-driven healthcare applications meet the highest ethical and clinical standards.

## Data Availability

All data were extracted from the referenced papers.

## Acknowledgments

The authors acknowledge the following manuscript reviewers: Michael Grasso, MD, PhD, FACP, FAMIA, ABPM-CI and Dipu Patel, DMSc, ABAIM, MPAS, PA-C

The authors utilized OpenAI’s Chat-GPT 4o for manuscript editing.

## Appendix A PRISMA-ScR Checklist

Appendix A – PRISMA-ScR Checklist
| SECTION | ITEM | PRISMA-ScR CHECKLIST ITEM | REPORTED ON PAGE # |
| --- | --- | --- | --- |
| **TITLE** |  |  |  |
| Title | 1 | Identify the report as a scoping review. | 1 |
| **ABSTRACT** |  |  |  |
| Structured summary | 2 | Provide a structured summary that includes (as applicable): background, objectives, eligibility criteria, sources of evidence, charting methods, results, and conclusions that relate to the review questions and objectives. | 1 |
| **INTRODUCTION** |  |  |  |
| Rationale | 3 | Describe the rationale for the review in the context of what is already known. Explain why the review questions/objectives lend themselves to a scoping review approach. | 2 |
| Objectives | 4 | Provide an explicit statement of the questions and objectives being addressed with reference to their key elements (e.g., population or participants, concepts, and context) or other relevant key elements used to conceptualize the review questions and/or objectives. | 3 |
| **METHODS** |  |  |  |
| Protocol and registration | 5 | Indicate whether a review protocol exists; state if and where it can be accessed (e.g., a Web address); and if available, provide registration information, including the registration number. | The protocol was not registered |
| Eligibility criteria | 6 | Specify characteristics of the sources of evidence used as eligibility criteria (e.g., years considered, language, and publication status), and provide a rationale. | 4 |
| Information sources* | 7 | Describe all information sources in the search (e.g., databases with dates of coverage and contact with authors to identify additional sources), as well as the date the most recent search was executed. | 5 |
| Search | 8 | Present the full electronic search strategy for at least 1 database, including any limits used, such that it could be repeated. | 5 |
| Selection of sources of evidence† | 9 | State the process for selecting sources of evidence (i.e., screening and eligibility) included in the scoping review. | 5 |
| Data charting process‡ | 10 | Describe the methods of charting data from the included sources of evidence (e.g., calibrated forms or forms that have been tested by the team before their use, and whether data charting was done independently or in duplicate) and any processes for obtaining and confirming data from investigators. | 6 |
| Data items | 11 | List and define all variables for which data were sought and any assumptions and simplifications made. | Appendix B |
| Critical appraisal of individual sources of evidence§ | 12 | If done, provide a rationale for conducting a critical appraisal of included sources of evidence; describe the methods used and how this information was used in any data synthesis (if appropriate). | Critical appraisal of included sources was not completed |
| Synthesis of results | 13 | Describe the methods of handling and summarizing the data that were charted. | 6 |
| **RESULTS** |  |  |  |
| Selection of sources of evidence | 14 | Give numbers of sources of evidence screened, assessed for eligibility, and included in the review, with reasons for exclusions at each stage, ideally using a flow diagram. | 8 |
| Characteristics of sources of evidence | 15 | For each source of evidence, present characteristics for which data were charted and provide the citations. | 7 |
| Critical appraisal within sources of evidence | 16 | If done, present data on critical appraisal of included sources of evidence (see item 12). | N/A |
| Results of individual sources of evidence | 17 | For each included source of evidence, present the relevant data that were charted that relate to the review questions and objectives. | 11 - 19 |
| Synthesis of results | 18 | Summarize and/or present the charting results as they relate to the review questions and objectives. | 11 - 19 |
| **DISCUSSION** |  |  |  |
| Summary of evidence | 19 | Summarize the main results (including an overview of concepts, themes, and types of evidence available), link to the review questions and objectives, and consider the relevance to key groups. | 20 |
| Limitations | 20 | Discuss the limitations of the scoping review process. | 23 |
| Conclusions | 21 | Provide a general interpretation of the results with respect to the review questions and objectives, as well as potential implications and/or next steps. | 24 |
| **FUNDING** |  |  |  |
| Funding | 22 | Describe sources of funding for the included sources of evidence, as well as sources of funding for the scoping review. Describe the role of the funders of the scoping review. | The study was not funded |

## Appendix B Data Extraction Tool

Appendix B – Data Extraction Tool
| Field | Description |
| --- | --- |
| Title | Full title as it appears in the publication. |
| Country of Study | Primary country where the study was conducted. |
| Study Start Date |  |
| Study End Date |  |
| Peer Review Status | Peer Reviewed / Not Peer Reviewed (Preprint Server) |
| Study Funding Sources | List all funding organizations or entities. |
| Possible Conflicts of Interest | Note any declared conflicts of interest by authors. |
| Study Aim | A concise statement of the study's primary objective. |
| Study Design | Specify the study design using standard types (e.g., RCT, Qualitative, Mixed-Methods, etc.). |
| Population Description | Describe the study population, including relevant demographics or patient characteristics. |
| Inclusion Criteria | Conditions that qualified participants for inclusion in the study. |
| Exclusion Criteria | Conditions or factors that excluded participants. |
| Type of RAG Model Description | Query-based, Latent Representation-based, Logit-based, etc. |
| Other type of RAG model | If selected, describe the model here. |
| Base Language Model | GPT, BERT, Google Gemini, Llama, or Other. |
| If GPT or Llama, specify version | e.g., 3.5, 4o, etc. |
| If other base model selected, list it here |  |
| Vector Database | Specify the database used for storing embeddings (e.g., Milvus, Weaviate). |
| Indexing Method | Describe the organization approach (e.g., IVF, HNSW). |
| Similarity Search Technique | Embedding generation, indexing, query processing, distance measurement, ANN search. |
| Retrieval Strategy | Top-K, Hybrid, Semantic Search, Keyword-based, ANN, etc. |
| Query Processing Method | Any preprocessing applied to queries to enhance retrieval accuracy. |
| Reranking Methods | Confidence-based, Semantic Similarity, BERT-based, LTR, RRF, etc. |
| Knowledge Base Sources | Medical journals, clinical guidelines, patient records, drug databases, etc. |
| Embedding Models | Domain-specific, general-purpose, or |
|  | other. |
| Model Training Details | Fine-tuning, zero-shot, few-shot. |
| Technology Stack | Open Source/Proprietary, Database Systems, Programming Languages, Frameworks, Cloud/On-Prem. |
| Intended User Group | Clinicians, students, administrators, etc. |
| Evaluation of RAG Effectiveness and Efficiency | Outcome measures, performance metrics, user satisfaction. |
| Health Informatics and Data Management | Privacy/security, compliance, data governance, interoperability. |
| Key Findings | Summary of main findings. |
| Intervention Description | Explain the RAG system implementation. |
| Comparison Type | Define any control or comparison groups. |
| Primary Outcomes | Main findings with statistical measures. |
| Secondary Outcomes | Additional findings or related measures. |
| Study Limitations | Describe limitations of the study. |
| Barriers to Implementation | Challenges in deployment. |
| Implications for Practice | Practical impact on healthcare or academic use. |
| Strategies to Overcome Implementation Challenges | Solutions to barriers. |
| User Training Requirements | Training provided for users. |
| Technical Support Provided | Ongoing support for system use. |
| User Interface Design Considerations | UX considerations in design. |
| Risk of AI Hallucination | Potential for generating incorrect data. |
| Ethical Issues Raised | Ethical concerns noted. |
| Bias Mitigation Strategies | Techniques to reduce bias. |
| Transparency and Explainability Measures | Steps to ensure explainability. |
| Handling of Large-Scale Data | Capacity for managing extensive data. |
| Response Time Metrics | System responsiveness and speed metrics. |
| Proposed Improvements | Potential system enhancements. |
| Potential New Applications | New uses in healthcare. |

